# Utilization of Genomic Services Among Physicians in Kenya

**DOI:** 10.1101/2025.02.04.25321649

**Authors:** Hillary Sigilai, Elijah Ogola, Syokau Ilovi

## Abstract

**Background:** With advancement of genetic technology and mainstreaming of genetics into all specialties of medicine, physicians will be required to play a pivotal role in the coordination and provision of primary genetic services.

**Objective:** To describe the delivery of basic genetic services among physicians in Kenya and factors affecting integration of genomic medicine into their practice.

**Materials and Methods:** An online descriptive cross-sectional study was conducted among registered physicians practicing within Kenya and physicians in training at University of Nairobi. A self-administered online close-ended questionnaire that assessed the following domains: delivery of genetic services, attitude and perceptions towards genetics, barriers to delivery of genomic services, knowledge of genetics and physician demographics. Simple random sampling method was used to recruit participants into the study. Interview of participants was conducted using an online close-ended questionnaire. Descriptive analysis methods were used for data analysis. The data was summarized using frequencies and proportions.

**Results:** The response rate was 41% with 190 of the eligible 464 physicians completing the survey. Eighty-seven percent of respondents had not received formal training in genetics, with 80% reporting involvement in evaluation of genetic patients. Physician involvement in genetic testing and pharmacogenomics was low at 31% and 29% respectively. Sixty-four percent of the respondents graded their graded their knowledge of genetics as moderate. Participants identified limited access to medical geneticist (80%), lack of referral guidelines (86%), high cost of genetic services (93%), inadequate knowledge of genetics as barriers to genetic service delivery.

**Conclusion:** Uptake of genetic service provision to patients by physician remains low due to inadequate genetics training, limited genetic specialists and prohibitive costs of genetic testing. Mitigation of these factors is required to improve genetic access to care.

## Background and Rationale

Genomic Medicine is a branch of medicine that deals with the prevention, diagnosis and management of genetic disorders in patients and their families. This encompasses both monogenic/Mendelian disorders and polygenic/complex trait/non-Mendelian disorders. As virtually all diseases are influenced or caused by the patients’ genetic architecture, it is imperative that genetics is incorporated into routine clinical care. With progression in the understanding the human genome, decreasing costs of genetic tests and widespread availability of testing, clinical genetics will continue to gain prominence in clinical care. Despite the clear benefits of clinical genetics in advancing human health, its uptake has remained low, with only about 50%-60% of physician providing genetic testing and counseling (1) (2) (3). Availability of critical infrastructure such as genetic testing remains limited, but their availability has started to increase. However, their applicability, availability, uptake and quality has not been examined. This study offers an overview of current trends in clinical genetics service utilization from the perspective of physicians. In Kenya, a physician is a specialist in Internal Medicine (standard 3- year postgraduate training). Findings from this study will aid in the formulation of institutional and departmental policies that will ensure effective utilization and delivery of genetic services in Kenya.

### Study Objective

To describe the delivery of basic genetic services among physicians and factors affecting integration of genomic medicine into their practice.

## Materials and Methods

### Study Design

Descriptive cross-sectional study using an anonymous self-administered online questionnaire.

### Study Population

This was a nationwide survey targeting physicians (specialists in internal medicine) in Kenya and Internal Medicine registrars at the University of Nairobi targeting to recruit 189 participants. The recruitment period ran from 7^th^ March 2024 to 29^th^ April 2024.

Inclusion criteria: Registered physicians practicing within Kenya and Internal Medicine registrars (physicians-in-training) at the University of Nairobi. Physicians and registrars who declined to give consent were excluded from the study.

Simple random sampling was used to recruit participants. Physicians’/registrars’ identifiers were obtained from the Kenya Medical and Practitioners’ and Dentists’ Council (KMPDC). A random seed was then set and executed to obtain appropriate numbers.

### Questionnaire design

Questionnaire used was adapted with permission from Carroll et al in 2012 in Canada (4). It was divided into 8 sections: Physicians’ demographic and characteristics (9 questions), current practice characteristics (10 items), family history taking (3 items), attitude towards genomic medicine (8 items), awareness of and experience with genetic services (9 items), knowledge of genetics (14 items) and barriers to delivery of genetic services (9itmes).

### Ethical considerations

Ethical approval was sought and obtained from Kenyatta National Hospital/University of Nairobi ethics review board (P859/11/2023). A written consent form in English language was provided describing the nature of the study, risks, potential benefits, their obligations, and the names and addresses of investigators to be contacted.

### Statistical analysis

Demographic characteristics were analyzed and presented as frequencies and percentages. Proportion of physicians who provide genetic services was presented as frequencies and percentage of physicians surveyed. Other answers were a mixture of 3-5 point Likert scales for knowledge, attitude, awareness and barriers. Each response was to be analysed individually as either positive or negative attitude and then scored as an aggregate score using the original Bloom’s cutoff. There was a total of 10 questions each with a score of 1- 5, i.e., a minimum of 10 and maximum of 50. Using Bloom’s cut off (42), attitude was categorized as positive /good >80% (40-50), neutral 60%-79%(30-39.5), and negative/poor <60%(<30 points).Knowledge domain was analysed as proportion using original Bloom’s cut off(42). A score <60% (< 27) being poor, >60%-79% (27-35.5 points) being moderate, and >80% (>36 points) being good knowledge.

The outcome variable, utilization of genetic services was defined as whether one had ordered carrier testing on parents of a child who was confirmed to have a genetic disorder and/or ordered a genetic test for genetic disorder. Factors associated with physicians’ use of genetic services; training in genetics, specialty, and number of years in practice were analysed using bivariate and multivariate logistic regression. Associations were reported by use of odds ratios and respective 95% confidence interval. P-values of <0.05 was considered as significant.

## Study Results

### Baseline Characteristics

The study recruited 190 participants from most of the sub-specialties on internal medicine. Majority (63%) had been in clinical practice for less than 10 years since completion of their basic undergraduate medical degree. Most of the respondents (87%) had not undertaken formal training in a genetics course (Table 1).

**Table 1:** Demographic characteristics of the participants.

| Variable | N = 190 |
| --- | --- |
| **Sex** |  |
| Female | 109 (57%) |
| Male | 81 (43%) |
| **Years in clinical practice** |  |
| 0-9 | 120(63%) |
| 10-19 | 56 (29%) |
| 20-29 | 8 (4.2%) |
| 30-39 | 6 (3.2%) |
| **Specialty** |  |
| Cardiology | 9 (4.7%) |
| Dermatology | 8 (4.2%) |
| Endocrinology | 9 (4.7%) |

| **Variable** | **N = 190** |
| --- | --- |
| Gastroenterology. | 1 (0.5%) |
| Infectious disease | 2 (1.1%) |
| Internal medicine (no sub-specialization) | 79 (42%) |
| Nephrology | 6 (3.2%) |
| Pulmonology | 2 (1.1%) |
| Registrar in internal medicine | 73 (38%) |
| Rheumatology | 1 (0.5%) |
| **Has training in genetics** (apart from embryology) | 25 (13%) |
| **Has a special interest in genetics** | 60 (32%) |

### Clinical Practice of Genetics

Eighty (80%) percent reported that evaluation and diagnosis of patients with genetic disorders was part of their practice, with 32% having ordered a genetic test and 24% referred a patient for specialist genetic evaluation. Only 14 (7%) respondents had performed carrier testing in parents of a child with a confirmed genetic disorder.

Majority (58%) of respondents complete a family history in over 75% of their patient populations, but only 48% updated the family history during subsequent clinic visits. Completion of a two or three generations family history was undertaken by 55% of respondents (Table 2).

**Table 2:** Family History taking.

| **Variable** | **N = 190** |
| --- | --- |
| **Proportion of patients you complete a family history** |  |
| 0 | 2 (1.1%) |
| 25% | 33 (17%) |
| 50% | 45 (24%) |
| 75% | 69 (36%) |
| 100% | 41 (22%) |
| **How often family history is updated** |  |
| At every visit | 20 (11%) |
| Every 2-4 years | 13 (6.8%) |
| Every 5-10 years | 17 (8.9%) |
| Never | 99 (52%) |
| Yearly or at periodic exam | 41 (22%) |
| **A two or three generation family history** | **104 (55%)** |
| **The family's ethnic background** | **116 (61%)** |

### Attitudes towards Genomic Medicine

The respondent’s attitude towards genomic medicine was scored in a Likert scale ranging from strongly disagree, to strongly agree. The responses were further categorized into three groups of negative attitudes (strongly disagree and disagree), neutral attitude (neutral), and positive attitude (strongly agree and agree).

The physicians surveyed in this study had an overall positive attitude towards genomics at 60% as illustrated below (Table 3).

**Table 3:** Physicians attitudes towards genomic medicine in categories.

| Category and scores (Bloom's cut off) | n | % | Mean (SD) |
| --- | --- | --- | --- |
| **Attitude** |  |  | 2.4 (0.4) |
| Positive Attitude (80% +) | 113 | 60% |  |
| Neutral (60-79%) | 60 | 31% |  |
| Negative Attitude (< 60%) | 17 | 19% |  |

Majority agreed that there is need to incorporate genomic medicine into their practice to improve patient outcomes. However, only 37% agreed that genomics was an exciting part of their practice. Furthermore, the majority agreed that genomic medicine is going to make an important contribution in the management of prenatal, paediatric, and adult conditions.

### Knowledge of Genetics Disorders

Overall, majority of respondents (64%) graded their knowledge of various aspects of genetics as moderate with only 7% having high level knowledge as illustrated below.

**Table 4:**
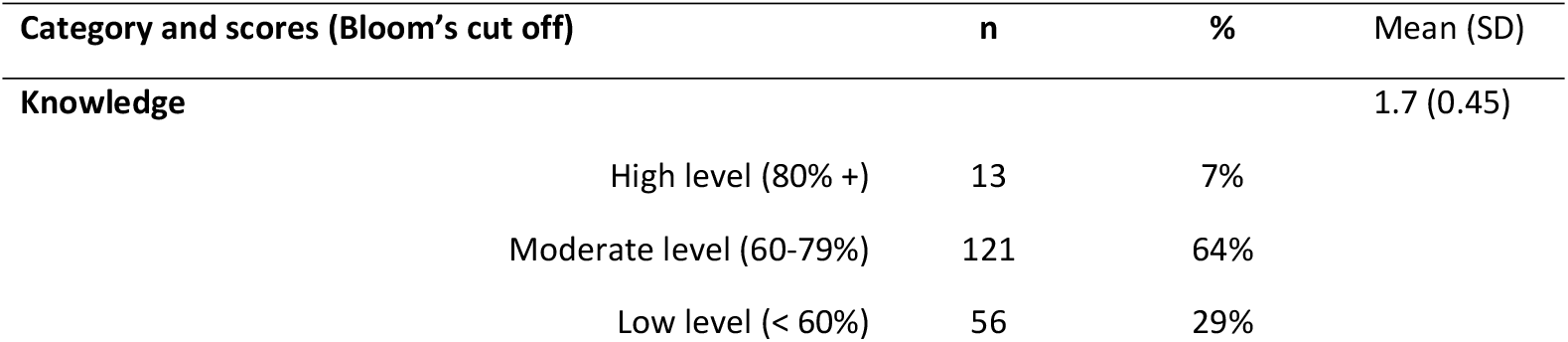
Physicians’ Self-perceived knowledge of genetics in categories.

Knowledge of emerging areas of genomic medicine such as newer technologies entering clinical practice (low-68%), interpreting results of ‘direct to consumer’ tests. (low-71%) was noticeably low. Majority (76%) do not know how to contact their local genetics centre or find information about genetic tests within the health system. (61%).

### Barriers to the provision of genetic services

Physicians identified limited access to medical geneticist (80%), lack of referral guidelines (86%), high cost of genetic services (93%), inadequate knowledge of genetics and limited access to genetic testing services as barriers to the provision of genetic services.

**Table 5:** Barriers to the provision of genetic services.

| **Variable** | **N = 190<sup>1</sup>** |
| --- | --- |
| **Limited access to a medical geneticist for referrals for testing and/or consultation** |  |
| Usually | 152 (80%) |
| **Lack of referral guidelines** |  |
| Usually | 163 (86%) |
| **High cost of genetic services such as testing.** |  |
| Usually | 177 (93%) |
| **Inadequate knowledge of genetics, genetic testing and genetic counselling.** |  |
| Usually | 117 (62%) |
| **Lack of patient interest in genetic evaluation.** |  |
| Usually | 78 (41%) |
| **Limited access to genetic testing services** |  |
| Usually | 155 (82%) |
| **Lack of detailed or updated family history** |  |

| Variable | N = 190<sup>1</sup> |
| --- | --- |
| Usually | 88 (46%) |

### Correlation between physician characteristics and their utilization of genetic services

There was a weak positive correlation between the years in clinical practice and the outcome variables above (p-value 0.018). Respondents who had training in genetics were also more likely to have ordered carrier testing on parents of a child who was confirmed to have a genetic disorder or ordered genetic test for a genetic disorder (p-value 0.037).

Years in clinical practice and whether one had received training were fit in the multivariate model. It was found that doctors who had more than 9 years of clinical practice were more likely to utilize genetic services compared to those with less than 9 years (adjusted OR = 1.16, 95% CI [1.16 – 4.09], p = 0.016]. Doctors who received training in genetics were more than twice likely to utilize genetic services compared to those who did not (aOR = 2.60, 95% CI [1.09- 6.29], p = 0.031).

## Discussion

The aim of this study was to offer the first comprehensive view of physicians’ involvement in genomic medicine, their awareness of currently available genetic services, attitudes towards genomic medicine and barriers to the integration of genomic medicine into mainstream healthcare in Kenya.

In this study majority of the participating physicians reported that they undertake clinical aspects of genomic medicine such as evaluation and diagnosis of patients with genetic disorders, referral of patients for specialised genetic services, and providing education about genetic conditions.

This indicates that basic genetic principles are incorporated in medical practice. Similar findings have been reported in other studies; from a Canadian survey of family physicians (5), Suchard et al(6);and Hayflick et al(3). However, involvement in areas such as genetic counselling and pharmacogenomics is low. It is debatable as to whether physicians should provide counselling to patients and families susceptible to inherited diseases. However, in the absence of professional genetic counsellors the role of physicians in counselling is integral. These findings are similar to those in literature with Haga et al(7) concluding in a survey of primary care physicians that only 13 % of respondents felt comfortable ordering pharmacogenomic tests while a quarter reported not having any education about pharmacogenetics. Capacity building and training is needed in these areas through continuous medical education as they are not incorporated into traditional medical education.

Worryingly, 68% of physicians have never ordered a genetic test for a genetic disorder. In addition, majority reported that they do not discuss benefits, risks, or limitations of genetic tests with patients or provide support to patients coping with genetic results. Even fewer know where to refer patients for genetic evaluation. The low uptake of genetic testing in our setting could be due to lack of training relevant to genetic testing, as well lack of knowledge and experience and limited testing infrastructure. The literature is mixed in this regard. In a 2008 survey of Primary care physicians in the US 60% of physicians had ordered a genetic test (1). Genetic testing has the potential decrease morbidity and mortality especially when used for risk profiling early in the management of chronic diseases (1).

Family history is a basic tool in clinical genetics and is key to risk assessment and identifying individuals who may benefit from further genetics evaluation(8). Key in family history taking is collecting adequate amounts of information and proper interpretation. 58% of physicians reported that they collect family history in 75%-100% of patients seen, with majority incorporating two or three generations (55%), family’s ethnic background (61%) and medical risk factors (93%). Previous surveys among physicians have shown an imbalance between frequency and quality of family history taking(5,9). Development of standardized family history taking tools with checklists and emphasis on proper history taking as part of medical education can be used to build on existing knowledge and skills in eliciting family history.

The study also looked at physicians’ attitude towards genomic medicine. The physicians surveyed in this study had a positive attitude towards genomic medicine at 60%. More than two- thirds of respondents felt that there is need to incorporate genomic medicine into their practice and keep up to date with advances in genomic medicine. Over half of the responding physicians agreed there are sufficient benefits to warrant testing for inherited adult-onset diseases and were convinced advances in genomic medicine will improve patient’s outcomes. However, majority do not find genetics and genomics an important part of their practice. This can be leveraged to enhance integration of genomic medicine into mainstream healthcare. In a south African study examining knowledge and attitudes towards predictive testing, attitude was largely positive despite the costs as they perceived benefits outweighed costs.(10) The results of this study also supports existing literature where participants felt that genomic medicine is likely to have an impact on clinical practice in future (5) (2,10).

Knowledge of genetics has been found to translate into increased uptake of genomic medicine into mainstream healthcare(10). Most (64%) of respondents rated their overall knowledge in genomic medicine as moderate i.e. 60-79% on bloom’s cutoff. Knowledge in basic concepts such as genetic consequences of consanguinity and genetic risk factors common complex disorders as moderate to high. However, knowledge of advanced/emerging themes such interpretation of direct-to-consumer testing and pharmacogenomics was largely poor. Specific educational interventions should be geared towards these areas. Self-perceived knowledge deficits are a global problem(11,12). More data is needed to determine specific educational resource needs and develop genetic educational programmes targeting physicians.

Systemic and individual barriers to providing genetic services have been previously identified in literature (13–15). In this study physicians identified limited access to medical geneticist (80%), lack of referral guidelines (86%), high cost of genetic services (93%), inadequate knowledge of genetics among others. These barriers impact practice, further impeding care delivery to patients in need of genetic services. Based on these findings multiple strategies can be deployed to ameliorate access to genetic services.

This needs assessment survey provides important insights into the state of genomic medicine in Kenya. The lack of awareness of existing genetic services and low uptake of genetic testing and counselling shows the need for investment in infrastructure such as genomic labs and training of personnel such as medical geneticists and genetic counsellors. There is need to develop genetics- training programmes and further incorporate genetics into the medical curriculum to improve the knowledge, skills, and confidence of physicians in genomic medicine.

## Conclusion and Recommendations

Study provides important insights into the state of genomic medicine in Kenya. Based on our findings, we recommend the following to improve the utilisation of genomic services among physicians in Kenya

i. Development of genetics training programmes and further incorporation of genetics into the medical curriculum.
ii. Investment in infrastructure such as genomic testing laboratories and training of personnel such as medical geneticists and genetic counsellors.

## Data Availability

All relevant data are available within the manuscript and its Supporting Information Files.

## Study Limitations

Possible response bias due to self-reporting. This was addressed by wording questions neutrally and ensuring answer options were open-ended.

Despite attaining the desired sample size of 189 overall response rate was low. The study lacked statistical power to analyse the secondary objective.

## Disclosures

This work was completed by H.S in part fulfilment of Master of Medicine in Internal Medicine from University of Nairobi

The authors did not receive funding for this work.

No artificial intelligence tools were used in production of this work.

